# Potassium-competitive acid channel blockers versus Proton-Pump inhibitors in the prevention of post-endoscopic peptic ulcer rebleeding: A systematic review and meta-analysis

**DOI:** 10.64898/2026.03.02.26346403

**Authors:** Nkengeh N. Tazinkeng, Shari Forbes, Richard McGowan, Miguel Agudelo, Mexan Mapouka, Bright C. Nwatamole, Sarpong Boateng, Cameg-Brenda Meriki, Smruti R. Mohanty

## Abstract

**Introduction:** Vonoprazan, a new oral potassium-competitive acid blocker (PCAB), has shown promise in terms of superior acid suppression when compared to Proton-pump inhibitors (PPIs). We evaluated the efficacy of PCABs versus PPIs in preventing rebleeding in high-risk peptic ulcer patients after endoscopic hemostasis.

**Methods:** Following the Preferred Reporting Items for Systematic Reviews and Meta-Analyses (PRISMA) guidelines, we conducted a comprehensive search for relevant studies across Medline, Embase, Web of Science, Cochrane Central Register of Controlled Trials and ClinicalTrials.gov, from inception till March 25, 2025. The primary outcome of interest was peptic ulcer rebleeding rate. Pooled risk ratios (RR) and mean difference (MD) with the corresponding 95% confidence intervals (CIs) were calculated.

**Results:** Three studies with 54,410 patients receiving endoscopic hemostasis for peptic ulcer bleeding were included in our analysis. The mean age of included participants was 71 ± 1.83 years.

There was no significant difference in rebleeding rates between patients receiving PPIs and PCABs (RR 0.827; 95 % CI: 0.5 –1.3). We observed a significant reduction in length of hospital stay in the PCAB group when compared to the PPI group (MD: -0.44, 95% CI: -0.72 - -0.17), but no significant difference in all-cause mortality between both groups (RR: 0.90, 95% CI: 0.79 - 1.04).

**Conclusions:** Our study demonstrates comparable efficacy of PPIs and PCABs in preventing rebleeding in patients with high-risk peptic ulcers after successful endoscopic hemostasis. However, there was a significant reduction in hospital length of stay favoring PCABs.

**Research in Context:** *What is already known on this topic:* Bleeding from peptic ulcers is considered one of the major reasons for mortality and hospitalization, and the standard treatment after endoscopic hemostasis is the administration of high-dose proton pump inhibitors (PPIs). Potassium competitive acid blockers (PCABs), such as vonoprazan, have been reported to have more potent and faster onset of action than PPIs in the treatment of acid-related diseases, but their efficacy in the prevention of post-endoscopic peptic ulcer rebleeding has not been well established in the literature in the form of a dedicated meta-analysis.

*What this study adds:* In the present study, the efficacy and safety of PCABs in the prevention of post-endoscopic rebleeding and mortality in 54,410 patients with high-risk peptic ulcer bleeding were investigated in the context of a systematic review and meta-analysis. PCABs were found to have similar efficacy to PPIs in the prevention of mortality and rebleeding in the context of endoscopic hemostasis, and the use of PCABs was also observed to reduce the length of stay in the hospital to a significant extent.

*How this study might affect research, practice or policy:* These findings indicate that PCABs are a reasonable alternative to PPIs in post-endoscopic management of high-risk peptic ulcer bleeding and may be particularly useful in situations where early discharge and optimization of resources are critical. Additional large-scale studies in different populations are required to validate these findings and create guidelines

## Introduction

Peptic ulcers are defects caused by gastric acid secretion or pepsin and typically extend to the muscularis propria layer of the gastric epithelium. They are commonly found in the stomach and proximal duodenum, however, they can also occur in the lower esophagus, distal duodenum or jejunum.(1) A manifestation of peptic ulcers is peptic ulcer bleeding (PUB), with an increasing proportion of bleeding episodes being related to non-steroidal anti-inflammatory drugs (NSAIDs) and aspirin use (2). Given PUB is a major cause of hospital admissions and mortality, it requires prompt endoscopic diagnosis and treatment.(3,4) Current guidelines recommend that patients with high-risk ulcers i.e., Forest 1A (active spurting), Forest 1B (active oozing) and Forest IIA (non-bleeding visible vessel) should undergo endoscopic hemostatic interventions (5). However, even with the advancement of these endoscopic interventions, rebleeding is of a significant concern, particularly in patients who have high-risk stigmata of bleeding, and bleeding from duodenal ulcers, including associated hemodynamic instability (6). High dose Proton-pump inhibitors (PPIs) have been the cornerstone of reducing these rebleeding episodes and are routinely administered prior to endoscopic hemostasis and after hemostasis for a total of 72 hours.(7,8) Studies have shown PPIs significantly reduce the rebleeding rate and mortality in patients resulting from its strong acid inhibition.(9) However, recently, Vonoprazan, a new oral potassium-competitive acid blocker with strong and sustained acid-inhibitory activity, has shown to have superior acid suppression when compared to PPIs.(10) It works by directly and competitively blocking the potassium binding site of the H+/K+ ATPase enzyme which causes a more rapid and sustained suppression of gastric acid secretion and unlike PPIs, it does not rely on acid activation (11). Vonoprazan has been shown to be superior to oral PPIs in the eradication of Helicobacter pylori (H. pylori) and has no inferiority with regards to the healing rate of erosive esophagitis.(12,13) In addition, a broader network meta-analysis showed that vonoprazan had higher ulcer healing and symptom remission rate compared to some PPI(14). It has also shown superiority in reducing delayed post-operative bleeding rate in ulcers caused by gastric endoscopic submucosal dissection.(15,16) In one study, *Hamada et al* conducted a single center randomised phase II trial comparing Vonoprazan and Lansoprazole.

They demonstrated that the bleeding rate in the Vonoprazon group was significantly lower than the threshold rate when compared to that of Lansoprazole in endoscopic submucosal dissection induced ulcers (11,17). Thus, it can be speculated that Vonoprazan has the potential to be more effective in reducing peptic ulcer rebleeding rates following endoscopic hemostasis, when compared to conventional intravenous PPIs. To date, there is no dedicated systematic review and meta-analysis that compare vonoprazan vs IV PPIs for peptic ulcer rebleeding after endoscopic therapy. Therefore, the aim of this systematic review and meta-analysis was to evaluate the efficacy of PCABs compared to PPIs in preventing rebleeding in high-risk peptic ulcer patients after successful endoscopic hemostasis.

## METHODS

We conducted this study following the updated Preferred Reporting Items for Systematic Reviews and Meta-Analyses (PRISMA) guidelines(18).

### Information Sources and Search Strategy

Electronic searches for published literature were conducted by a medical librarian [RM] using Medline, Embase, Web of Science, Cochrane Central Register of Controlled Trials and ClinicalTrials.gov, from inception until March 25, 2025. The search strategy contained three keywords, linked using the AND operator:

1. (Vonoprazan OR “potassium-competitive acid blocker*” OR “P-CAB”)
2. (“Proton pump inhibitor” OR “Proton pump inhibitors” OR “Proton pump inhibitors”[Pharmacological Action] OR “Proton pump inhibitors”[MeSH Terms] OR Omeprazole OR Esomeprazole OR Lansoprazole OR Dexlansoprazole OR Pantoprazole OR Rabeprazole)
3. (ulcer OR “peptic ulcer”[MeSH Terms] OR “peptic ulcer” OR “high risk ulcer” OR “bleeding” OR “rebleeding” OR “hemorrhage”[MeSH Terms] OR “hemorrhage” OR “hemorrhaging”)

### Inclusion and Exclusion Criteria

Studies were eligible for inclusion if: (1) they were randomized controlled studies, retrospective or prospective observational studies; (2) they had full-text available; (3) they compared the efficacy of P-CABs vs PPIs in preventing peptic ulcer rebleeding. Review articles, letters, conference proceedings, case series, and case report studies were excluded. Studies not published in English and without quantitative data of interest were also excluded.

### Data Extraction and Management

All identified studies from the search strategy were de-duplicated using EndNote (EndNote, 2013)(19) and then uploaded into the Covidence systematic review software (Covidence, 2024)(20).

Titles, abstracts, and keywords of all the articles were screened by two independent reviewers, and irrelevant reports were removed. Full-text screening of the selected articles was also performed by two reviewers independently and cross-checked for eligible studies. Discrepancies were resolved through team discussion or adjudicated by a third reviewer (NT). The following data were extracted from each included study: author name, year of publication, country, study design, sample size, duration of follow up, mean age, sex, ulcer location, forest classification, type, dose, route and duration of PCAB and PPI, and length of follow-up. The outcomes of interest included rebleeding rates, all-cause mortality and hospital length of stay. We used the definitions of rebleeding that were provided by each study (Table 1).

**Table 1:** Characteristics of included studies.

| Study | Country | Study design | Sample size (PCAB:PPI) | PPI regimen | PCAB regimen | Follow up duration | Outcomes assessed | Rebleeding definition |
| --- | --- | --- | --- | --- | --- | --- | --- | --- |
| Yabe et al., 2024* | Japan | Retrospective (PSM) | 27098:27098 | daily doses of<br>Lansoprazole 30 mg,<br>Rabeprazole 10 mg,<br>Esomeprazole 20 mg and<br>Omeprazole 20 mg | Vonoprazan 20 mg daily | NA | Rebleeding rate, In-hospital mortality | overt bleeding that required endoscopic hemostasis and/or transfusion at 2 days after the initial endoscopic hemostasis |
| Pattarapuntakul et al., 2024 | Thailand | RCT | 9:11 | First 72 h: pantoprazole infusion at 8 mg per hour for 72 h and oral placebo every 12 h<br>Day 3-14: oral omeprazole at 20 mg every 12 h, along with an oral placebo every 12 h<br>Day 15 - 56: oral omeprazole at 20 mg once daily, along with an oral placebo once daily | First 72 h: oral vonoprazan at a dose of 20 mg every 12 h and a placebo infusion for 72 h<br>Day 3-14: Oral vonoprazan at 20 mg every 12 h, in addition to oral placebo every 12 h<br>Day 15 - 56: Oral vonoprazan at 20 mg once daily, along with an oral placebo, from day 15 to day 56 | 56 days | Rebleeding rates (3 days, 30 days), Complete ulcer healing rate | Presence of recurrent hematemesis, passage of melena, and/or fresh blood staining in the nasogastric tube after documented successful endoscopic hemostasis; in addition to any of the following criteria: (i) hypotension (SBP< 90 mm Hg) or tachycardia (HR> 120/min) without other explained causes; (ii) a decrease in HB> 2 g/dL during any 24 h period; or (iii) receiving at least 2 units of blood transfusion within 72 h |
| Seeratragool et al., 2024 | Thailand | RCT | 98:96 | Day 0-3: intravenous PPI at the rate of 8 mg/h<br>Day 4-28: oral omeprazole 20 mg twice daily | Day 0-3: Oral Vonoprazan 20 mg twice daily,<br>Day 4-28: Oral Vonoprazan 20 mg once daily for 28 days | 30 days | Rebleeding rates (3, 7 and 30 days), 30-day mortality rate, blood transfusion requirement, rescue therapy needed for recurrent bleeding, and length of hospital stay | recurrent hematemesis with significant amounts of fresh blood (>200 mL) or fresh blood in the nasogastric tube aspirate, and/or hematochezia or melena after stool color normalization, hypotension (SBP <90 mm Hg), or tachycardia (HR>100/min) with melena, or a decrease in hemoglobin by more than 2 g/dL with melena |

PSM: propensity score matching, RCT: Randomized control trial, SBP: Systolic Blood Pressure, HR: Heart Rate

### Risk of Bias Assessment

We used the Risk of Bias tool for randomized trials (RoB2) to assess the quality of randomized control trials, and the “Risk Of Bias In Non-randomised Studies - of Interventions” (ROBINS-I) tool to assess the quality of non-randomized observational studies (21,22). RoB2 addresses 5 domains namely; bias due to randomization, bias due to deviations from intended interventions, bias due to missing outcome data, bias in measurement of outcomes, and bias in selection of the reported result. The domains ROBINS-I addresses are; bias due to confounding, bias in classification of interventions, bias in selection of participants into the study, bias due to deviations from intended interventions, bias due to missing data, bias in measurement of outcomes, and bias in selection of the reported result.

## Statistical Analysis

All statistical analyses were conducted using the Comprehensive Meta-Analysis (CMA) 3.0 software. A random-effects model was employed to calculate the pooled risk ratio (RR) and standardized mean difference (MD) with corresponding 95% Confidence Interval (95%CI). The random-effects model was chosen as it accounts for potential between-study heterogeneity, providing a more conservative estimate of the pooled treatment effect. Statistical significance was defined as p<0.05. Heterogeneity among studies was assessed using Cochran’s Q test, where p<0.10 was considered indicative of significant heterogeneity. The I^2^ statistic was used to quantify heterogeneity, with values interpreted as follows: 30 to 50% represents moderate heterogeneity, 50 to 75% represents substantial heterogeneity, and >75% represents considerable heterogeneity. The presence of small-study effects was not assessed using a funnel plot, and Egger’s test for funnel plot asymmetry was not performed, as each outcome included fewer than 10 studies, making these assessments unreliable

## RESULTS

Of the 779 studies retrieved from database searches, 97 were identified as duplicates and removed. After screening titles and abstracts, and assessing full-text articles for eligibility, three studies met the inclusion criteria and were included in our meta-analysis. The search process and selection results are shown in Fig. 1 Two of the three included studies were randomized control trials conducted in Thailand and one study was observational in design (propensity-matched retrospective cohort study) and published from Japan. In total, studies evaluated 54,410 patients receiving endoscopic hemostasis for peptic ulcer bleeding - 27,205 patients receiving post-endoscopy prescription of P-CAB and 27,205 receiving post-endoscopy prescription of PPI. The mean age of participants across all three studies was 71 (±1.76) years. In all three studies, Vonoprazan was the main P-CAB drug used and was offered to participants at a dose of 20 mg twice daily via oral route. On the other hand, the most common PPI prescribed was Omeprazole at a dose 20 mg twice daily via oral route. (Table 1)

**Figure 1.**
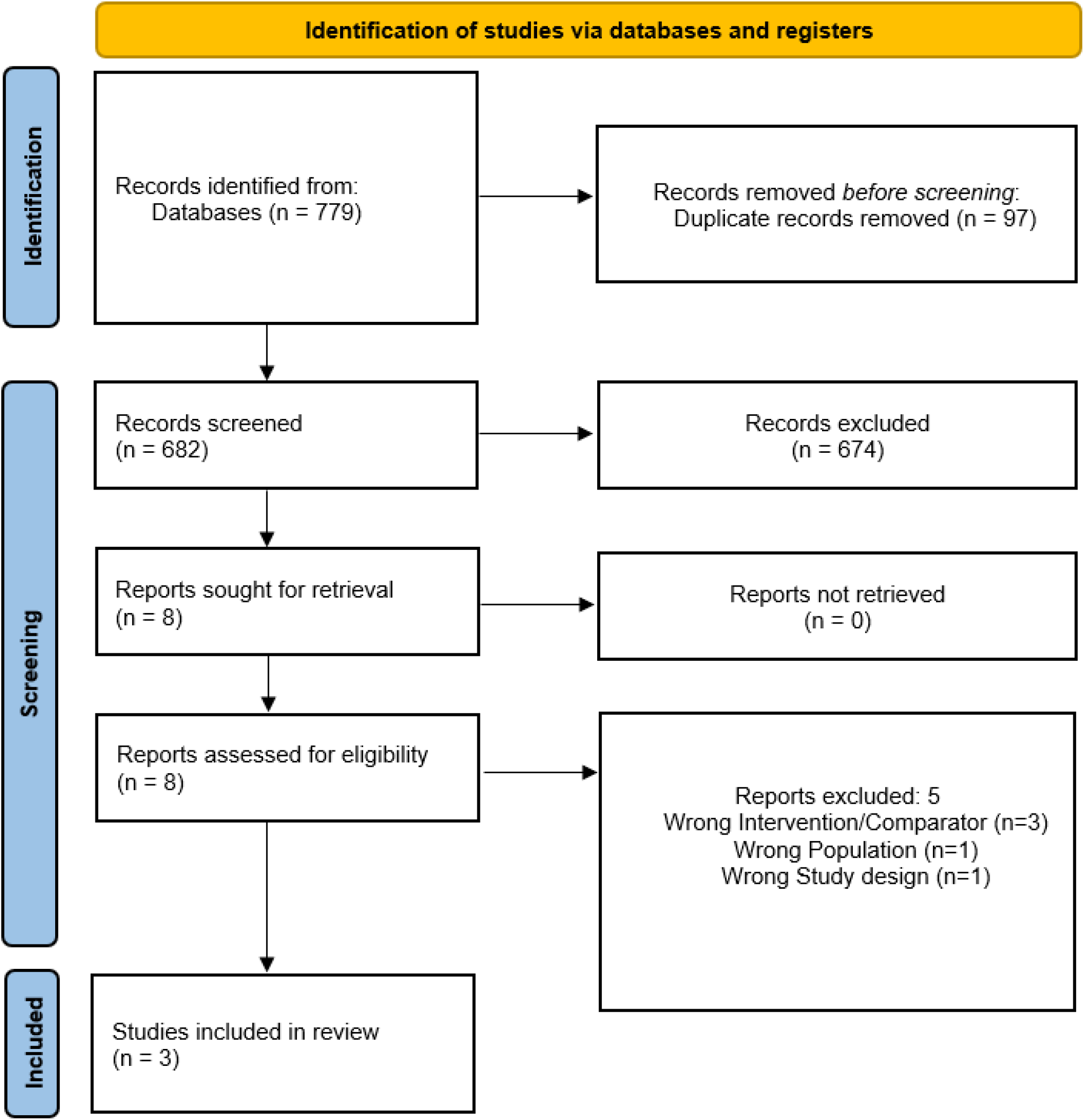
PRISMA Flow Diagram of Study Selection.

**Figure 2.**
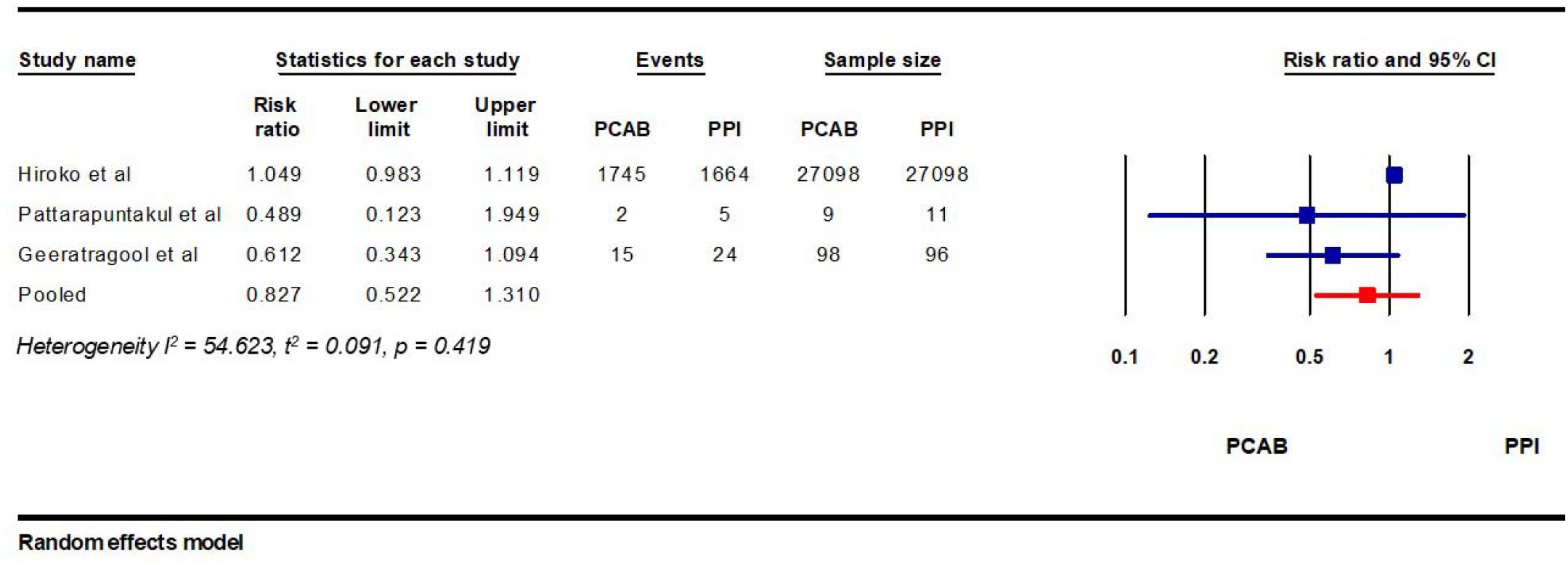
Relative Risk of rebleeding in patients receiving PPI vs PCAB post endoscopic hemostasis.

**Figure 3.**
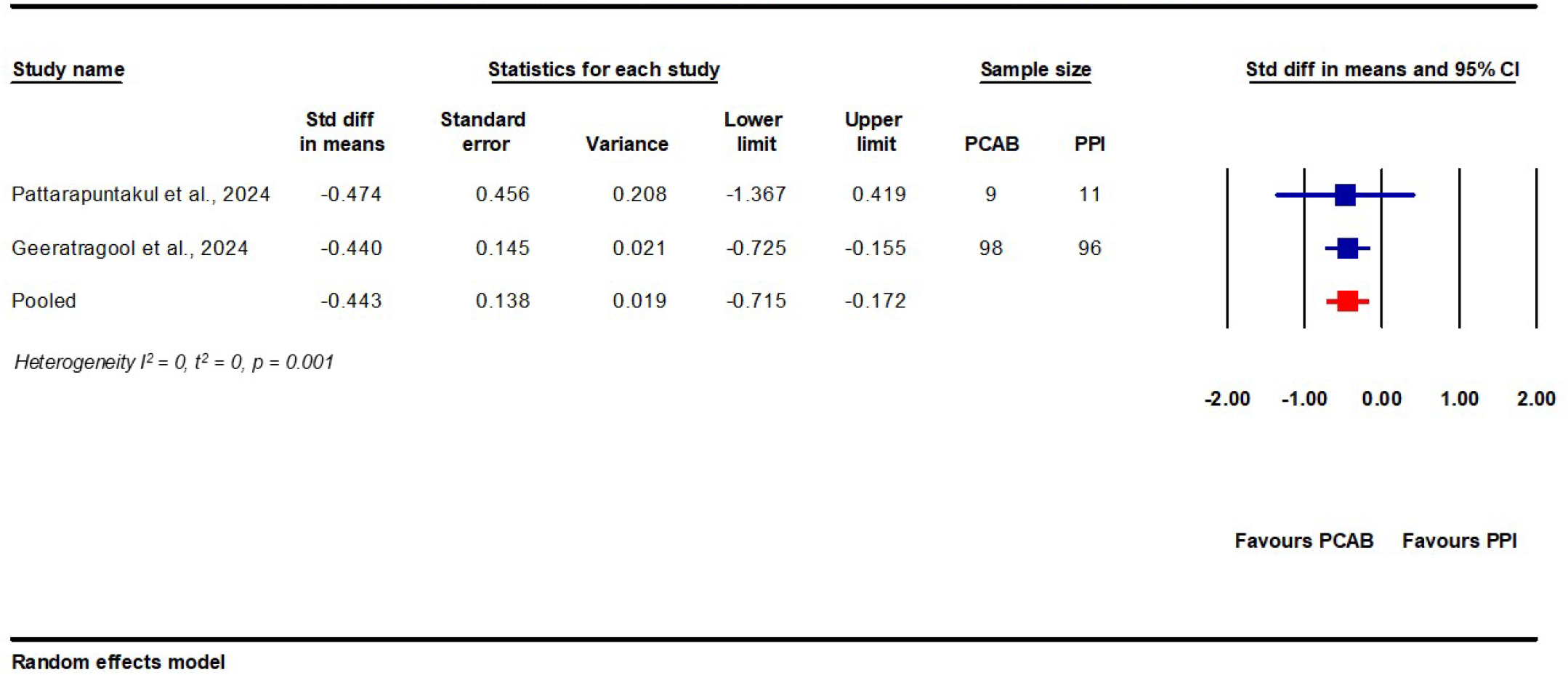
Length of hospital stay in patients receiving PPI vs PCAB after endoscopic hemostasis.

**Figure 4.**
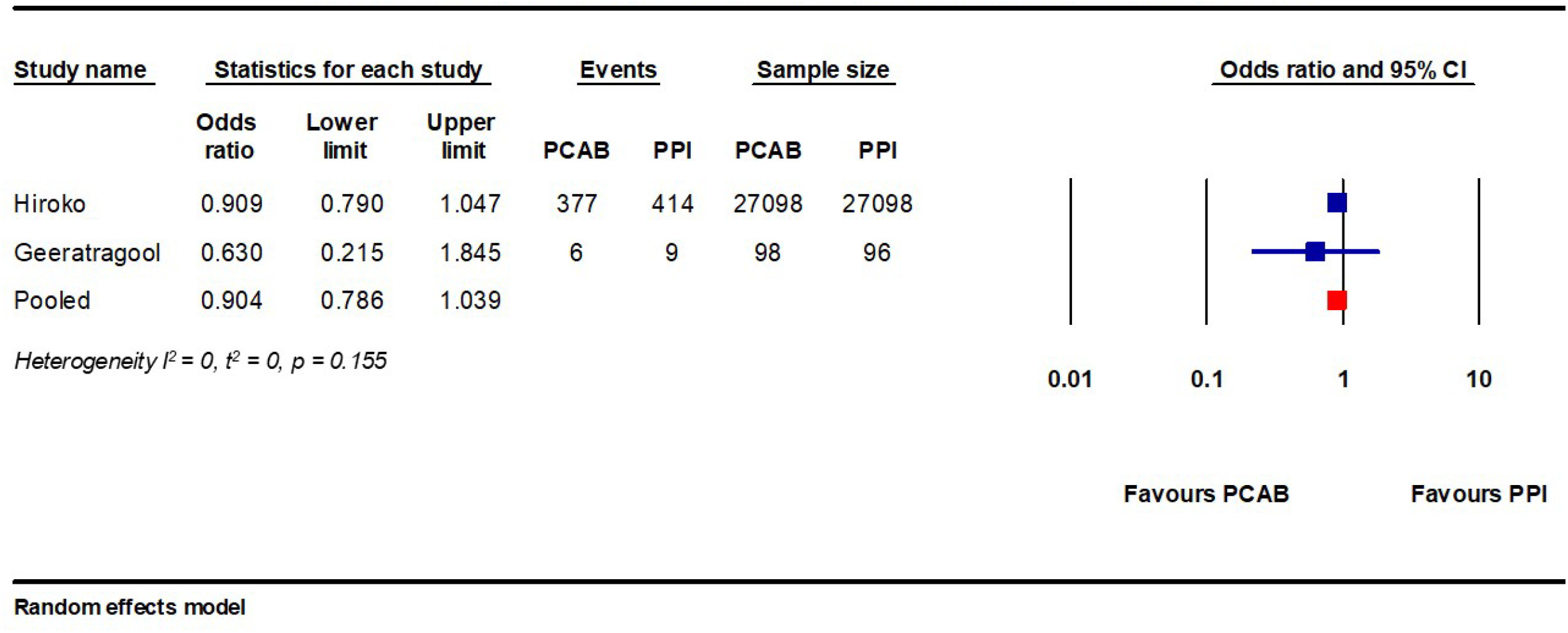
All-cause mortality in patients receiving PPI vs PCAB after endoscopic hemostasis.

### Meta-analysis outcomes

In our pooled analysis, 1693 patients had peptic ulcer rebleeding in the PPI group and 1762 patients had rebleeding in the PCAB group (6.2% vs 6.5%). On risk analysis, there was no significant difference in risk of rebleeding between both groups of patients (RR 0.827; 95 % CI: 0.5 –1.3, *p*= 0.419, I^2^ = 54.6%)

In terms of length of hospital stay, we observed a significant reduction in length of hospital stay in the PCAB group when compared to the PPI group. Specifically, PCAB use after endoscopic hemostasis was associated with a mean reduction in length of hospital stay of 0.44 days (95% CI: 0.17–0.72 days, *p*= 0.001), indicating that patients in the PCAB group were discharged approximately half a day earlier on average. The consistency of the treatment effect across studies was notable, with an I^2^ value of 0 %, suggesting no significant heterogeneity.

We found no significant difference in all-cause mortality between patients receiving PPI and PCAB following endoscopic hemostasis (1.6% vs 1.4%; RR: 0.90, 95% CI: 0.79 - 1.04, *p* = 0.155, I^2^ = 0).

## Quality assessment

The outcomes of the RoB2 and ROBINS-I evaluation for risk of bias are presented in *Supplementary Figure 1*. Studies that met our inclusion criteria were deemed to have moderate risk of bias. There were no studies with serious or critical risk of bias, which would have prompted exclusion from our meta-analysis. Specifically, *Abe et al. (2024)* was judged to be at **moderate risk of bias**, as a result of residual confounding from unmeasured bleeding severity and exclusion of patients with missing data. Using the RoB2 tool for randomized studies, the study by *Geeratragool et al (2024)* was judged to have moderate risk of bias, primarily due to its open-label non-inferiority design, whilst the study by *Pattarapuntakul T*. et al *(2024)* was judged to have moderate risk of bias owing to per-protocol-only analysis.

## Discussion

Peptic ulcer bleeding is a major cause of hospital admissions and carry a significant morbidity and mortality, especially in those with high-risk stigmata or multiple comorbid conditions(23); Standard practice to achieve hemostasis and promote healing involves a combination of endoscopic treatment followed by high dose PPI for 72 hours after intervention(8). Despite advancement in care, 10-15% of patients will rebleed within 30 days, with the highest incidence in the first 7 days(24), which is associated with a two-to fivefold increase in mortality (25).

In this systematic review and meta analysis of studies evaluating the efficacy of PCABs compared to PPIs for prevention of peptic ulcer rebleeding after endoscopic hemostasis, we included three studies encompassing more than 54,000 participants. We demonstrated that there was no significant difference in peptic ulcer rebleeding and mortality between PCABs vs PPIs. However, there was a significant reduction in length of hospital stay by approximately half a day with PCAB use.

Our study highlights the fact that despite the observed pharmacological advantage of PCAB in ensuring rapid and more stable and sustained acid suppression, this advantage does not translate into reduced rebleeding rates. As suggested by the European Society of Gastrointestinal Endoscopy (ESGE) and American Gastroenterological Association (AGA) guidelines, patients with peptic ulcer bleeding usually receive empiric treatment with high dose intravenous PPI before endoscopy, as PPIs have been shown to promote clot stability. (26–28) This pre-endoscopic administration of PPI could effectively raise gastric pH above 4, a critical threshold for healing that makes it hard to observe any significant advantage of superior drugs administered following endoscopic hemostasis.(29) Another possible explanation for the observed similarity in rebleeding rates between PPIs and PCABs is the fact that if endoscopic hemostasis is effectively achieved, this could minimize the effects of further acid suppression. Our finding therefore suggests that other factors such as ulcer characteristics, comorbidities or endoscopic factors may play a significant role in prevention of rebleeding than just post-procedural acid suppression alone.

The significant reduction in length of hospital stay in patients receiving PCABs could be a reflection of their faster and more sustained acid suppression, ensuring earlier clinical stability and faster transition to oral intake. Although the reduction in length of hospital stay was modest, it is still possible that it may play a significant role in reducing healthcare utilization and costs, particularly in resource-limited health settings.

We did not find any significant difference in all-cause mortality between both groups. This might be a reflection of the relatively low mortality rates observed across included studies. In addition many cases of mortality following peptic ulcer bleeding are usually driven by factors such as advanced age, comorbidities and complications not usually related to rebleeding.(30)

Our study adds to the existing pool of studies that have shown non-inferiority of PCABs compared to PPI in the treatment of acid-related gastrointestinal diseases. It adds to existing literature by specifically highlighting the similarity in clinical outcomes between both classes of acid suppressants in the post-endoscopic setting.

This study is the first systematic review and meta-analysis that compared PCABs to PPIs in preventing peptic ulcer rebleeding in patients post-endoscopic hemostasis. The use of a random-effect model, and an independent dual reviewer process strengthens the reliability of our findings.

Our study has several limitations. All studies were conducted exclusively in the Asian populations, limiting their generalizability to other populations where there are significant differences in population characteristics, prescribing patterns and clinical practice guidelines. In addition, only three studies met our inclusion criteria, thereby limiting our ability to perform subgroup analysis.

Although baseline demographics were similar across both groups, clinical factors such as ulcer severity, concomitant antithrombotic use, and long-term outcomes were limited or inconsistently reported across studies, potentially resulting in biased pooled estimates. These limitations highlight the need for larger, multicenter randomized trials across diverse settings to further clarify the role of newer acid suppressants such as PCABs in preventing peptic ulcer rebleeding after endoscopic hemostatasis. These studies should also include stratified analysis based on ulcer characteristics and bleeding risk to identify potential subgroups that might benefit most from PCAB therapy.

In conclusion, this study demonstrates that PCAB has comparable efficacy to PPIs in preventing peptic ulcer rebleeding and mortality after endoscopic hemostasis. However, PCAB was associated with reduction in length of hospital stay. These findings suggest that in clinical settings where early discharge and optimization of healthcare resources are prioritized, PCABs are a reasonable alternative to PPIs in the post-endoscopic management of peptic ulcer bleeding.

## Data Availability

The datasets during and/or analyzed during the current study are available from the corresponding author on reasonable request.

## Graphical Abstract Text

In this systematic review and meta-analysis of 54,410 patients with high-risk peptic ulcer bleeding treated endoscopically, potassium-competitive acid blockers (PCABs) showed similar rates of rebleeding and mortality when compared with proton pump inhibitors (PPIs). However, using PCABs was linked to a significantly shorter hospital stay. This suggests they are comparable in effectiveness, with possible benefits for healthcare use.

## Notes

**Conflict of Interest Disclosure:** The authors declare no conflict of interest

### Competing Interest Statement

The authors have declared no competing interest.

### Funding Statement

The was no funding for this project

## REFERENCES

1. Malik TF, Gnanapandithan K, Singh K. Peptic Ulcer Disease. In: StatPearls [Internet]. Treasure Island (FL): StatPearls Publishing; 2025 [cited 2025 Dec 28]. Available from: http://www.ncbi.nlm.nih.gov/books/NBK534792/

2. Sung JJY, Tsoi KKF, Ma TKW, Yung MY, Lau JYW, Chiu PWY. Causes of mortality in patients with peptic ulcer bleeding: a prospective cohort study of 10,428 cases. Am J Gastroenterol. 2010 Jan;105(1):84–9.

3. Kim JS, Park SM, Kim BW. Endoscopic Management of Peptic Ulcer Bleeding. Clin Endosc. 2015 Mar;48(2):106–11.

4. Holster IL, Kuipers EJ. Update on the Endoscopic Management of Peptic Ulcer Bleeding. Curr Gastroenterol Rep. 2011;13(6):525–31.

5. Yen HH, Wu PY, Wu TL, Huang SP, Chen YY, Chen MF, et al. Forrest Classification for Bleeding Peptic Ulcer: A New Look at the Old Endoscopic Classification. Diagn Basel Switz. 2022 Apr 24;12(5):1066.

6. Vakil N. Endoscopic Diagnosis, Grading, and Treatment of Bleeding Peptic Ulcer Disease. Gastrointest Endosc Clin N Am. 2024 Apr;34(2):217–29.

7. Khan MA, Howden CW. The Role of Proton Pump Inhibitors in the Management of Upper Gastrointestinal Disorders. Gastroenterol Hepatol. 2018 Mar;14(3):169–75.

8. Laine L, Barkun AN, Saltzman JR, Martel M, Leontiadis GI. ACG Clinical Guideline: Upper Gastrointestinal and Ulcer Bleeding. Am J Gastroenterol. 2021 May 1;116(5):899–917.

9. Rockey DC. Proton pump inhibitors in acute peptic ulcer bleeding. Gastroenterology. 2005 Aug 1;129(2):756–7.

10. Oshima T, Miwa H. Potent Potassium-competitive Acid Blockers: A New Era for the Treatment of Acid-related Diseases. J Neurogastroenterol Motil. 2018 July;24(3):334–44.

11. Padwale V, Kirnake V, Daswani R, Kodmalwar A, Gupta A. A Comprehensive Review on the Efficacy and Safety of Vonoprazan in the Management of Gastric Acid-Related Diseases. Cureus. 2024 July;16(7):e64777.

12. Laine L, DeVault K, Katz P, Mitev S, Lowe J, Hunt B, et al. Vonoprazan Versus Lansoprazole for Healing and Maintenance of Healing of Erosive Esophagitis: A Randomized Trial. Gastroenterology. 2023 Jan 1;164(1):61–71.

13. Ozercan M, Tawheed A, Ismail A, Amer MS, El-Kassas M. Vonoprazan and proton pump inhibitors: Which is superior for Helicobacter pylori eradication? World J Gastroenterol. 2025 May 7;31(17):103156.

14. Ouyang M, Zou S, Cheng Q, Shi X, Zhao Y, Sun M. Comparative Efficacy and Safety of Potassium-Competitive Acid Blockers vs. Proton Pump Inhibitors for Peptic Ulcer with or without Helicobacter pylori Infection: A Systematic Review and Network Meta-Analysis. Pharmaceuticals. 2024 June;17(6):698.

15. Hidaka Y, Imai T, Inaba T, Kagawa T, Omae K, Tanaka S. Efficacy of vonoprazan against bleeding from endoscopic submucosal dissection-induced gastric ulcers under antithrombotic medication: A cross-design synthesis of randomized and observational studies. PLOS ONE. 2021 Dec 23;16(12):e0261703.

16. Chen L, Jiang D, Hu D, Cui X. Comparison of vonoprazan and proton pump inhibitors for the treatment of gastric endoscopic submucosal dissection-induced ulcer: an updated systematic review and meta-analysis. BMC Gastroenterol. 2024 Mar 15;24(1):110.

17. Hamada K, Uedo N, Tonai Y, Arao M, Suzuki S, Iwatsubo T, et al. Efficacy of vonoprazan in prevention of bleeding from endoscopic submucosal dissection-induced gastric ulcers: a prospective randomized phase II study. J Gastroenterol. 2019 Feb;54(2):122–30.

18. Page MJ, McKenzie JE, Bossuyt PM, Boutron I, Hoffmann TC, Mulrow CD, et al. The PRISMA 2020 statement: an updated guideline for reporting systematic reviews. BMJ. 2021 Mar 29;372:n71.

19. The EndNote Team. EndNote. 2013.

20. Covidence [Internet]. [cited 2025 June 29]. Covidence - Better systematic review management. Available from: https://www.covidence.org/

21. Sterne JAC, Savović J, Page MJ, Elbers RG, Blencowe NS, Boutron I, et al. RoB 2: a revised tool for assessing risk of bias in randomised trials. 2019 Aug 28 [cited 2025 July 22]; Available from: https://www.bmj.com/content/366/bmj.l4898.short?casa_token=vV-xBsDq6HQAAAAA:c_UYYw6mOOJAJ-Pt9BCXzGrisVMGeQTKmdWqR9WHFottSrATx92GZWerjDjPsCJ4eGXw-bjVjJg

22. Sterne JA, Hernán MA, Reeves BC, Savović J, Berkman ND, Viswanathan M, et al. ROBINS-I: a tool for assessing risk of bias in non-randomised studies of interventions. 2016 Oct 12 [cited 2025 July 22]; Available from: https://www.bmj.com/CONTENT/355/BMJ.I4919.abstract

23. Chiu PWY, Ng EKW, Cheung FKY, Chan FKL, Leung WK, Wu JCY, et al. Predicting mortality in patients with bleeding peptic ulcers after therapeutic endoscopy. Clin Gastroenterol Hepatol Off Clin Pract J Am Gastroenterol Assoc. 2009 Mar;7(3):311–6; quiz 253.

24. Rosenstock SJ, Møller MH, Larsson H, Johnsen SP, Madsen AH, Bendix J, et al. Improving quality of care in peptic ulcer bleeding: nationwide cohort study of 13,498 consecutive patients in the Danish Clinical Register of Emergency Surgery. Am J Gastroenterol. 2013 Sept;108(9):1449–57.

25. Rockall TA, Logan RF, Devlin HB, Northfield TC. Risk assessment after acute upper gastrointestinal haemorrhage. Gut. 1996 Mar;38(3):316–21.

26. Mullady DK, Wang AY, Waschke KA. AGA Clinical Practice Update on Endoscopic Therapies for Non-Variceal Upper Gastrointestinal Bleeding: Expert Review. Gastroenterology. 2020 Sept 1;159(3):1120–8.

27. Xiao X, Liu X, Yan H, Xing X, Luo X, Yang J. Proton pump inhibitor therapy after transcatheter angiography in refractory nonvariceal acute upper gastrointestinal bleeding patients: a cohort study. BMC Gastroenterol. 2024 May 17;24(1):168.

28. Gralnek IM, Stanley AJ, Morris AJ, Camus M, Lau J, Lanas A, et al. Endoscopic diagnosis and management of nonvariceal upper gastrointestinal hemorrhage (NVUGIH): European Society of Gastrointestinal Endoscopy (ESGE) Guideline - Update 2021. Endoscopy. 2021 Mar;53(3):300–32.

29. van Rensburg CJ, Cheer S. Pantoprazole for the Treatment of Peptic Ulcer Bleeding and Prevention of Rebleeding. Clin Med Insights Gastroenterol. 2012 Sept 17;5:51–60.

30. Sung JJY, Tsoi KKF, Ma TKW, Yung MY, Lau JYW, Chiu PWY. Causes of mortality in patients with peptic ulcer bleeding: a prospective cohort study of 10,428 cases. Am J Gastroenterol. 2010 Jan;105(1):84–9.

